# Multi-site clinical validation of Isothermal Amplification based SARS-COV-2 detection assays using different sampling strategies

**DOI:** 10.1101/2021.07.01.21259879

**Authors:** Kanan T. Desai, Karla Alfaro, Laura Mendoza, Matthew Faron, Brian Mesich, Mauricio Maza, Rhina Dominguez, Adriana Valenzuela, Chyntia DÍaz Acosta, Magaly MartÍnez, Juan C. Felix, Rachel Masch, Sofia Gabrilovich, Michael Plump, Akiva P. Novetsky, Mark H. Einstein, Nataki C. Douglas, Miriam Cremer, Nicolas Wentzensen

**Author notes:** **Corresponding author** Kanan T. Desai, Clinical Genetics Branch, Division of Cancer Epidemiology and Genetics, National Cancer Institute, NIH, USA.

## Abstract

**Background:** Isothermal amplification-based tests were developed as rapid, low-cost, and simple alternatives to real-time reverse transcriptase-polymerase chain reaction (RT-PCR) tests for SARS-COV-2 detection.

**Methods:** Clinical performance of two isothermal amplification-based tests (Atila Biosystems iAMP COVID-19 detection test and OptiGene COVID-19 Direct Plus RT-LAMP test) was compared to clinical RT-PCR assays using different sampling strategies. A total of 1378 participants were tested across four study sites.

**Results:** Compared to standard of care RT-PCR testing, the overall sensitivity and specificity of the Atila iAMP test for detection of SARS-CoV-2 were 76.2% and 94.9%, respectively, and increased to 88.8% and 89.5%, respectively, after exclusion of an outlier study site. Sensitivity varied based on the anatomic collected site. Sensitivity for nasopharyngeal was 65.4% (range across study sites:52.8%-79.8%), mid-turbinate 88.2%, saliva 55.1% (range across study sites:42.9%-77.8%) and anterior nares 66.7% (range across study sites:63.6%-76.5%). The specificity for these anatomic collection sites ranged from 96.7% to 100%. Sensitivity improved in symptomatic patients (overall 82.7%) and those with a higher viral load (overall 92.4% for ct≤25). Sensitivity and specificity of the OptiGene Direct Plus RT-LAMP test, conducted at a single study-site, were 25.5% and 100%, respectively.

**Conclusions:** The Atila iAMP COVID test with mid-turbinate sampling is a rapid, low-cost assay for detecting SARS-COV-2, especially in symptomatic patients and those with a high viral load, and could be used to reduce the risk of SARS-COV-2 transmission in clinical settings. Variation of performance between study sites highlights the need for site-specific clinical validation of these assays before clinical adoption.

## Introduction

The COVID-19 (Coronavirus Disease of 2019) pandemic has led to major disruptions in health services worldwide. In many developed nations, widespread SARS-CoV-2 (Severe acute respiratory syndrome coronavirus 2) testing and mass vaccination has allowed for a return to most elective health services. However, many low- and middle-income countries (LMICs) have limited access to testing and vaccination and continue to struggle to contain COVID-19 (1) (2). As the COVID-19 crisis continues, considerable reductions of cancer screening, cancer control, and elective clinical services remain (3). The safe return to cancer screening and elective testing and procedures during the pandemic, especially in low vaccination regions, requires reliable SARS-CoV-2 testing for both providers and patients.

Numerous SARS-CoV-2 detection assays have been developed and introduced into the market under emergency use authorizations (EUA) (4). EUAs are granted primarily based on analytic sensitivity (i.e., Limit of Detection (LOD)) and analytic specificity (i.e., cross-reactivity) with limited clinical validations. Yet, a thorough clinical performance evaluation of SARS-CoV-2 assays in important to understand the strengths, limitations, and specific applications of these assays (5). Current Centers for Disease Control (CDC) guidelines recommend the use of laboratory-based nucleic acid amplification test (NAAT) (e.g., reverse trasnscriptase-polymerase chain reaction (RT-PCR)) for confirmatory testing. Specimens that are considered optimal for detection include nasopharyngeal (NP), nasal mid-turbinate, and anterior nasal swabs. Currently, the CDC does not recommend NAAT that use oral specimens (e.g., saliva) for confirmatory testing (6–8).

In addition to clinical performance, several other factors are important to consider when assessing feasibility of an assay for use in different environments and clinical settings. These factors include time to run the assay, hands-on time, throughput, ease of implementation, and cost. Furthermore, the possibility to use different sampling approaches, including self-collection, can be an important distinguishing feature since many LMICs have limited personal protective equipment (PPE). While RT-PCR assays fulfill the desired clinical performance criteria, they are not ideal for primary care clinics in resource-limited settings as point-of-care SARS-COV-2 screening tests due to high costs as well as longer turn around times and need for technical expertise (9). While rapid antigen-based tests address these limitations, they lack sensitivity to rule out an active infection (10). Isothermal amplification-based reverse transcription assays may fill this gap as they are more rapid (take ∼ 1 hour), cheaper (∼5-15 USD per test) and simpler (not needing RNA extraction) than RT-PCR based tests (11), but require clinical validation.

The primary objective of this study was to evaluate the clinical performance and operational characteristics of two isothermal amplification-based SARS-CoV-2 tests: 1) iAMP COVID-19 detection test (Atila BioSystems, USA) targeting N and ORF1a-genes of SARS-COV-2 virus, and 2) COVID-19 Direct Plus RT-LAMP test (OptiGene Ltd., UK) targeting ORF1ab-gene of SARS-COV-2 virus, compared to clinical RT-PCR tests. The secondary objective was to evaluate the influence of different sampling strategies on the detection of SARS-COV-2. One specific use for such assays is rapid SARS-COV-2 testing to allow for a safer return to preventive clinical encounters such as cancer screening in low- and middle-income countries.

## Materials and Methods

### Study design and population

A cross-sectional study was conducted from December 2020 to April 2021 at four clinical sites: Hospital Nacional de Santa Ana, El Salvador; Hospital Materno Infantil de San Lorenzo, Ministerio de Salud Pública (MSP-BS), Paraguay; Medical College of Wisconsin (MCW), USA; and Rutgers New Jersey Medical School (NJMS), USA (Table 1). The study protocol and sampling strategies varied slightly across the study sites, based on local requirements.

**Table 1:**
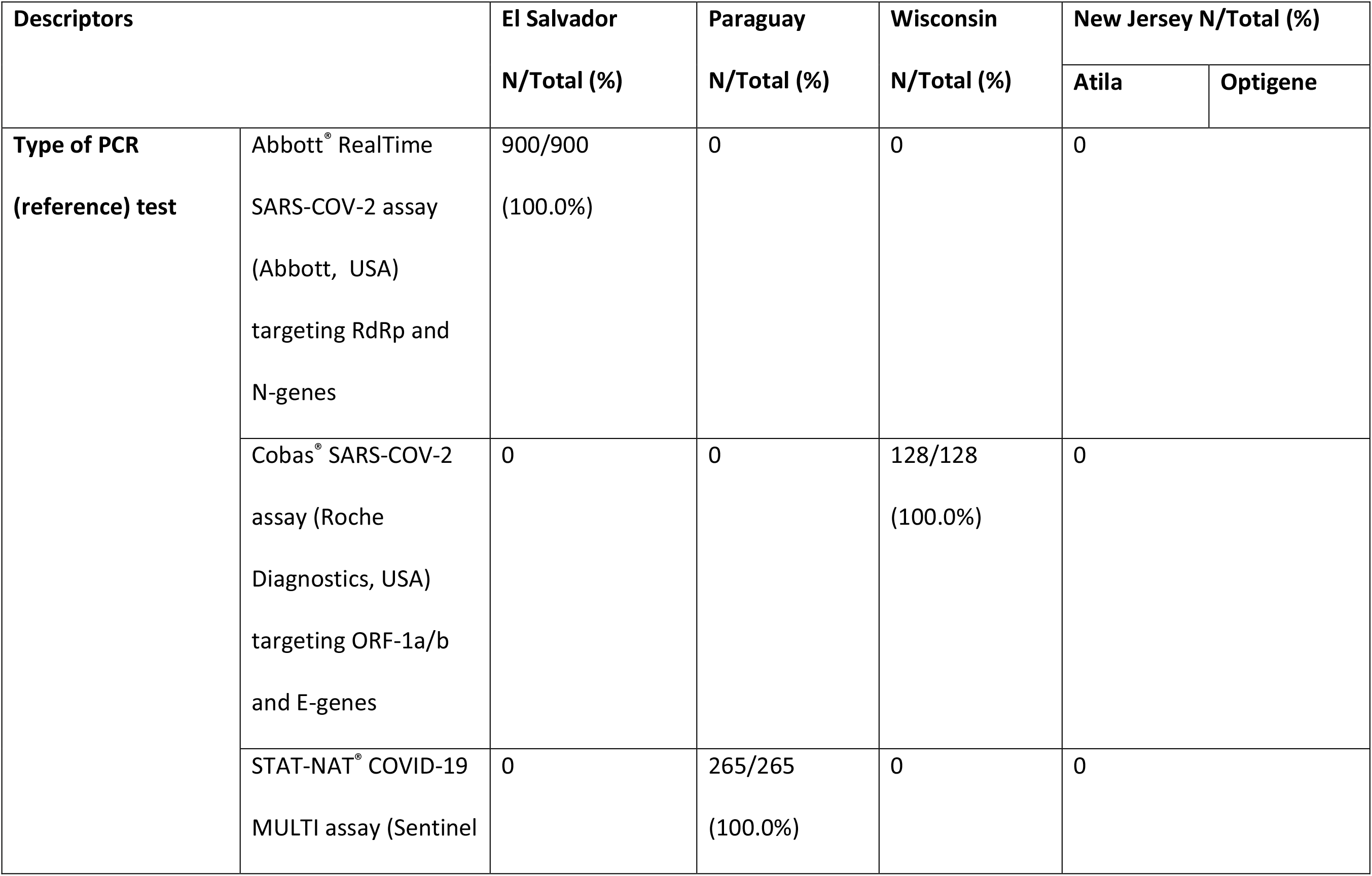
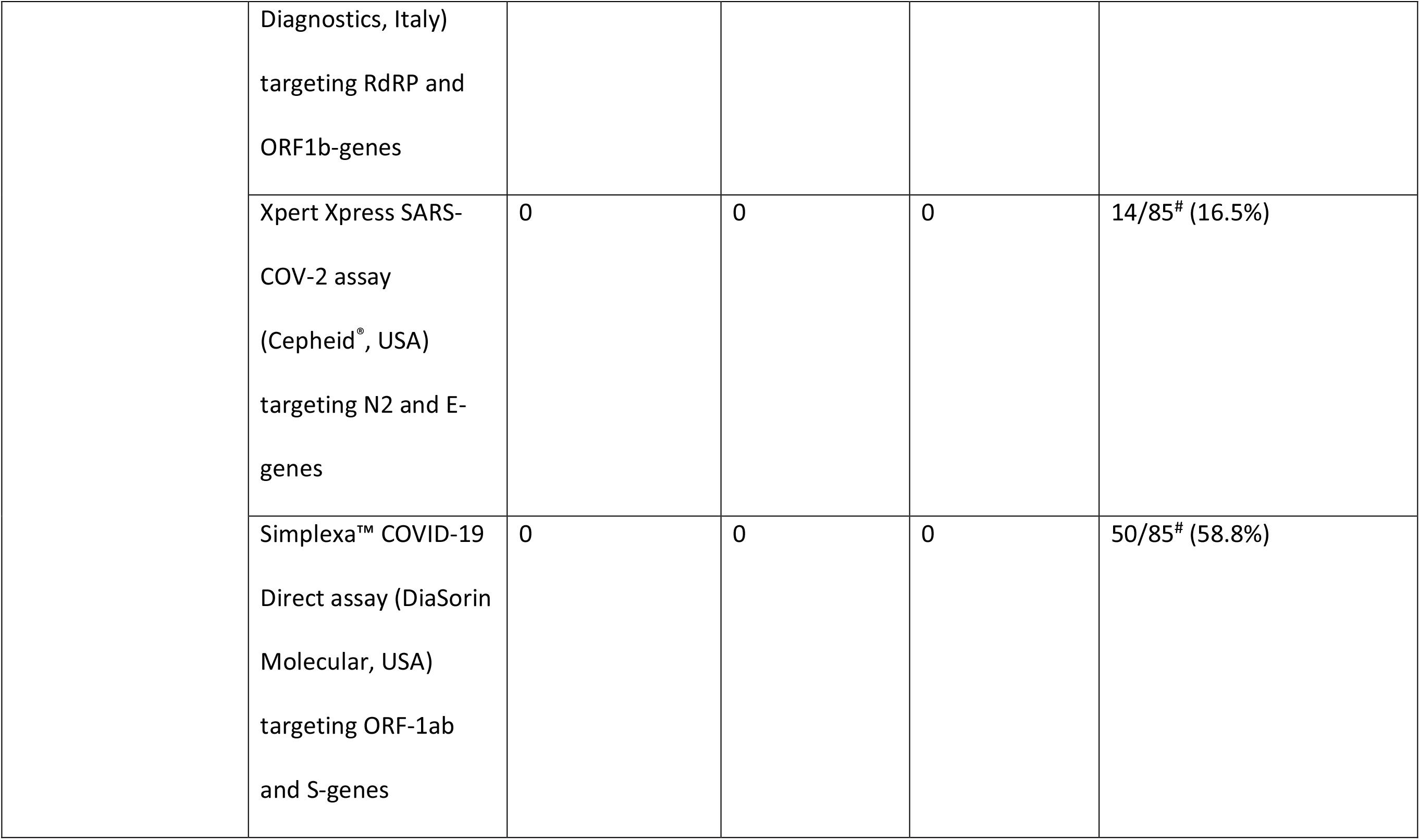
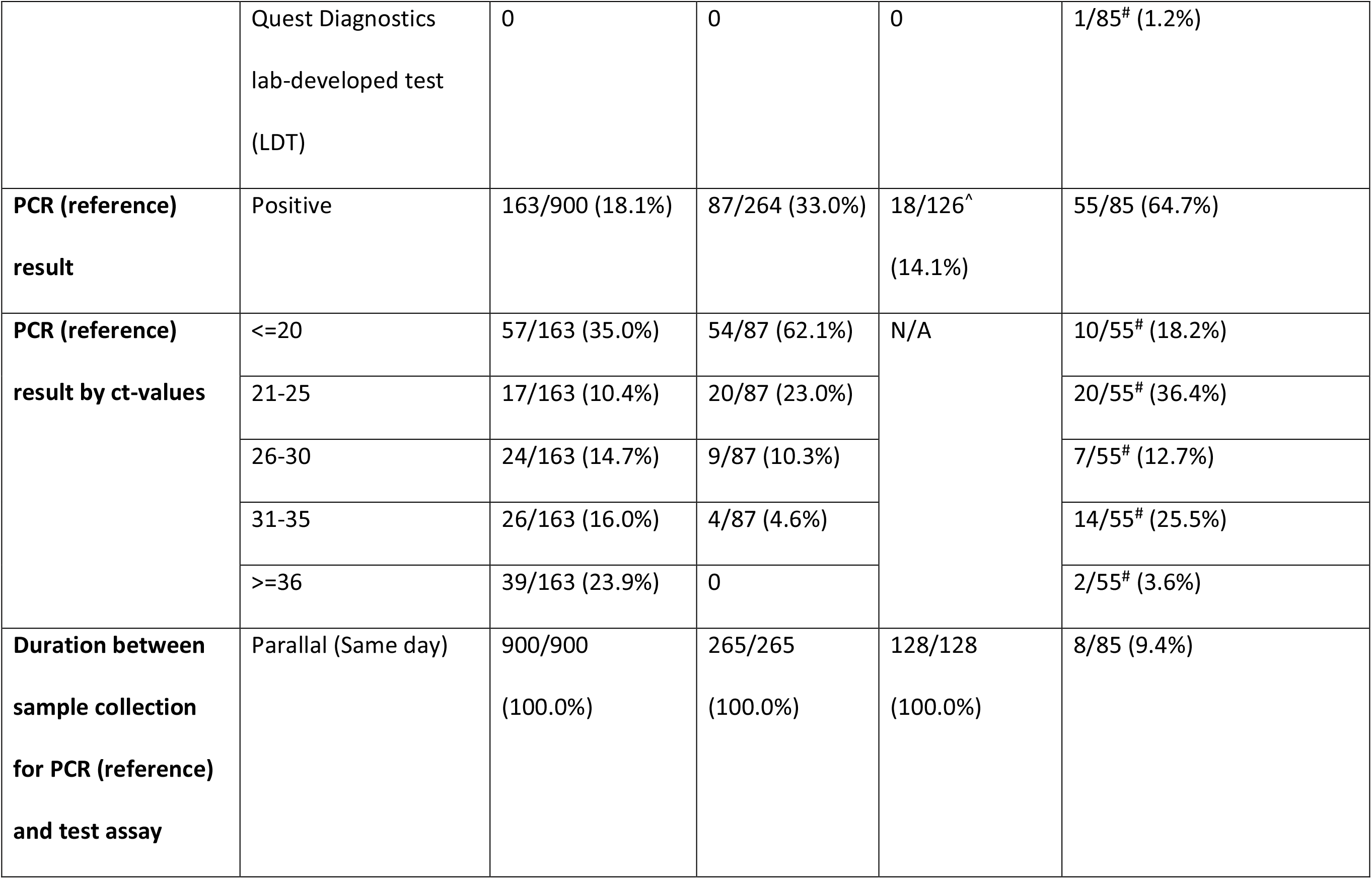
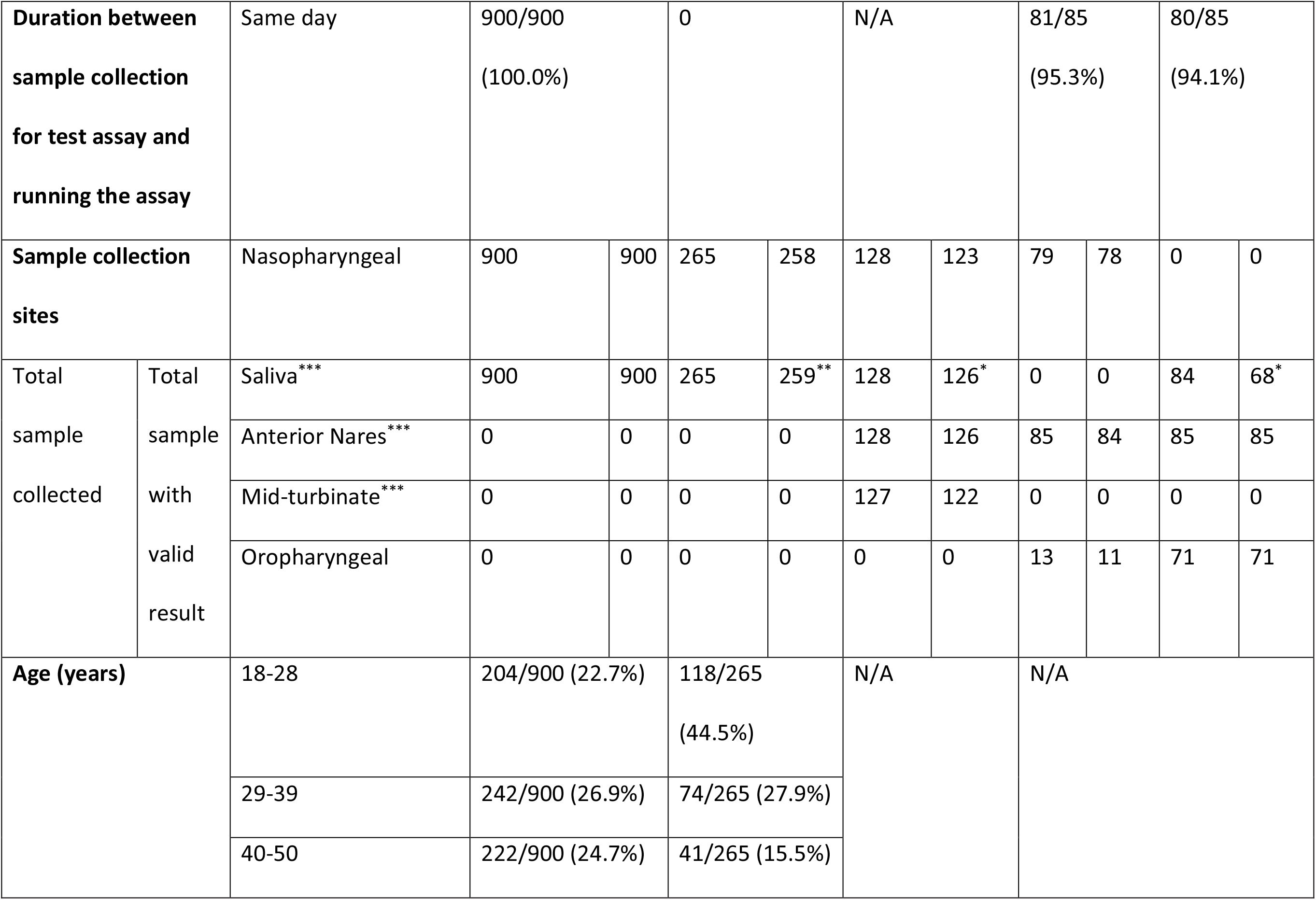
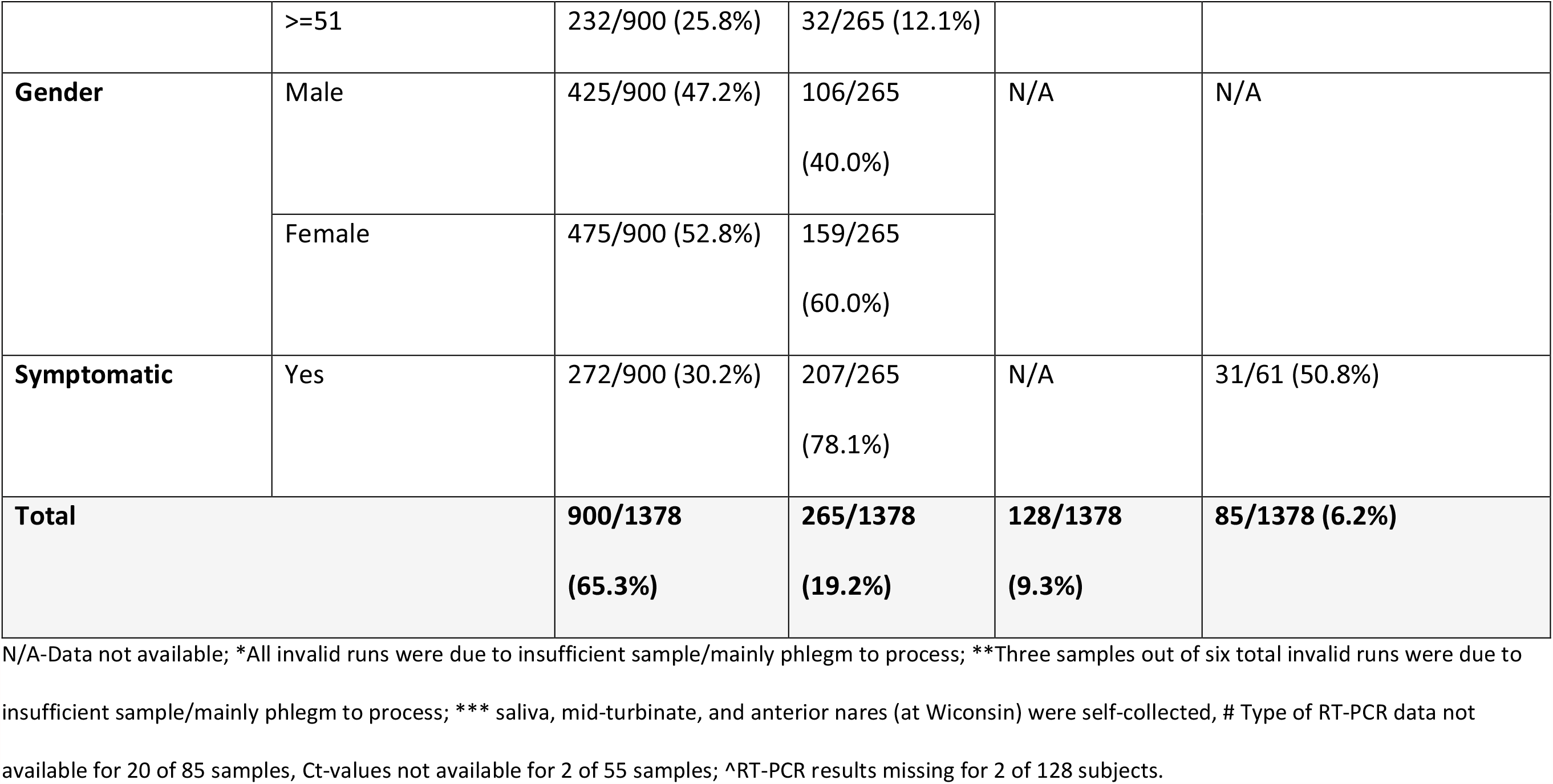
Description of the study population.

| Descriptors |  | El Salvador<br>N/Total (%) | Paraguay<br>N/Total (%) | Wisconsin<br>N/Total (%) | New Jersey N/Total (%) |  |
| --- | --- | --- | --- | --- | --- | --- |
|  |  |  |  |  | Atila | Optigene |
| **Type of PCR**<br>**(reference) test** | Abbott® RealTime<br>SARS-COV-2 assay<br>(Abbott, USA)<br>targeting RdRp and<br>N-genes | 900/900<br>(100.0%) | 0 | 0 | 0 |  |
|  | Cobas® SARS-COV-2<br>assay (Roche<br>Diagnostics, USA)<br>targeting ORF-1a/b<br>and E-genes | 0 | 0 | 128/128<br>(100.0%) | 0 |  |
|  | STAT-NAT® COVID-19<br>MULTI assay (Sentinel | 0 | 265/265<br>(100.0%) | 0 | 0 |  |

|  |  |  |  |  |  |
| --- | --- | --- | --- | --- | --- |
|  | Diagnostics, Italy)<br>targeting RdRP and<br>ORF1b-genes |  |  |  |  |
|  | Xpert Xpress SARS-COV-2 assay<br>(Cepheid®, USA)<br>targeting N2 and E-genes | 0 | 0 | 0 | 14/85<sup>#</sup> (16.5%) |
|  | Simplexa™ COVID-19<br>Direct assay (DiaSorin<br>Molecular, USA)<br>targeting ORF-1ab<br>and S-genes | 0 | 0 | 0 | 50/85<sup>#</sup> (58.8%) |
|  | Quest Diagnostics<br>lab-developed test<br>(LDT) | 0 | 0 | 0 | 1/85<sup>#</sup> (1.2%) |
| **PCR (reference)**<br>**result** | Positive | 163/900 (18.1%) | 87/264 (33.0%) | 18/126<sup>^</sup><br>(14.1%) | 55/85 (64.7%) |
| **PCR (reference)**<br>**result by ct-values** | <=20 | 57/163 (35.0%) | 54/87 (62.1%) | N/A | 10/55<sup>#</sup> (18.2%) |
|  | 21-25 | 17/163 (10.4%) | 20/87 (23.0%) |  | 20/55<sup>#</sup> (36.4%) |
|  | 26-30 | 24/163 (14.7%) | 9/87 (10.3%) |  | 7/55<sup>#</sup> (12.7%) |
|  | 31-35 | 26/163 (16.0%) | 4/87 (4.6%) |  | 14/55<sup>#</sup> (25.5%) |
|  | >=36 | 39/163 (23.9%) | 0 |  | 2/55<sup>#</sup> (3.6%) |
| **Duration between**<br>**sample collection**<br>**for PCR (reference)**<br>**and test assay** | Parallal (Same day) | 900/900<br>(100.0%) | 265/265<br>(100.0%) | 128/128<br>(100.0%) | 8/85 (9.4%) |

|  |  |  |  |  |  |  |  |  |  |  |  |  |
| --- | --- | --- | --- | --- | --- | --- | --- | --- | --- | --- | --- | --- |
| Duration between sample collection for test assay and running the assay |  | Same day | 900/900<br>(100.0%) |  | 0 |  | N/A |  | 81/85<br>(95.3%) |  | 80/85<br>(94.1%) |  |
| Sample collection sites |  | Nasopharyngeal | 900 | 900 | 265 | 258 | 128 | 123 | 79 | 78 | 0 | 0 |
| Total sample collected | Total sample with valid result | Saliva*** | 900 | 900 | 265 | 259** | 128 | 126* | 0 | 0 | 84 | 68* |
|  |  | Anterior Nares*** | 0 | 0 | 0 | 0 | 128 | 126 | 85 | 84 | 85 | 85 |
|  |  | Mid-turbinate*** | 0 | 0 | 0 | 0 | 127 | 122 | 0 | 0 | 0 | 0 |
|  |  | Oropharyngeal | 0 | 0 | 0 | 0 | 0 | 0 | 13 | 11 | 71 | 71 |
| Age (years) |  | 18-28 | 204/900 (22.7%) |  | 118/265<br>(44.5%) |  | N/A |  | N/A |  |  |  |
|  |  | 29-39 | 242/900 (26.9%) |  | 74/265 (27.9%) |  |  |  |  |  |  |  |
|  |  | 40-50 | 222/900 (24.7%) |  | 41/265 (15.5%) |  |  |  |  |  |  |  |

|  |  |  |  |  |  |
| --- | --- | --- | --- | --- | --- |
|  | >=51 | 232/900 (25.8%) | 32/265 (12.1%) |  |  |
| **Gender** | Male | 425/900 (47.2%) | 106/265<br>(40.0%) | N/A | N/A |
|  | Female | 475/900 (52.8%) | 159/265<br>(60.0%) |  |  |
| **Symptomatic** | Yes | 272/900 (30.2%) | 207/265<br>(78.1%) | N/A | 31/61 (50.8%) |
| **Total** |  | **900/1378**<br>**(65.3%)** | **265/1378**<br>**(19.2%)** | **128/1378**<br>**(9.3%)** | **85/1378 (6.2%)** |

N/A-Data not available; \*All invalid runs were due to insufficient sample/mainly phlegm to process; \*\*Three samples out of six total invalid runs were due to insufficient sample/mainly phlegm to process; \*\*\* saliva, mid-turbinate, and anterior nares (at Wiconsin) were self-collected, # Type of RT-PCR data not available for 20 of 85 samples, Ct-values not available for 2 of 55 samples; ^RT-PCR results missing for 2 of 128 subjects.

At the El Salvador site, 900 asymptomatic and symptomatic subjects presenting for SARS-COV-2 testing were enrolled. A standard NP swab for RT-PCR was collected from all the participants for clinical diagnosis. A second provider-collected dry NP swab and a self-collected direct saliva sample were obtained from study participants in parallel for the Atila iAMP test.

At the Paraguay site, 265 asymptomatic and symptomatic subjects presenting for SARS-COV-2 testing were enrolled in the study. A standard NP swab for RT-PCR was collected from all the participants for clinical diagnosis. In addition, for those consenting to participate in the study, a leftover of the clinical NP swab placed in viral transport medium (VTM) and a second parallel self-collected direct saliva sample were obtained for the Atila iAMP test.

At the Wisconsin (MCW) site, 128 symptomatic and asymptomatic subjects presenting for SARS-COV-2 testing were enrolled in the study. A standard NP swab for RT-PCR was collected from all the participants for clinical diagnosis. In addition, for those consenting to participate, a second provider-collected dry NP swab, a self-collected dry mid-turbinate swab, a self-collected dry anterior nares swab, and a self-collected direct saliva sample were obtained in parallel for Atila iAMP test.

At the New Jersey (NJMS) site, 55 symptomatic SARS-COV-2 positive patients, based on a prior RT-PCR assay, who were admitted for observation and management of COVID-19 were enrolled in the study. 28 of 55 (50.9%) of the patients were enrolled within 24 hours, 14 of 55 (25.5%) within 48 hours, and 6 of 55 (10.9%) within 72 hours of the sample collection for the RT-PCR test. In addition, 30 participants expected to be negative for the SARS-COV-2 infection (i.e., no SARS-COV-2 symptoms) were enrolled. A negative SARS-COV-2 RT-PCR test obtained within five days of test sample collection was performed on 28 (93.3%) of these 30 participants. Regardless of the RT-PCR status, for everyone enrolled in the study, a provider-collected dry NP swab and a provider-collected dry anterior nares swab was obtained at the time of enrollment for the Atila iAMP test. In addition, a provider-collected oropharyngeal (OP) swab placed in Sigma Virocult^®^medium (MSW, UK) and a self-collected direct saliva sample was also obtained at the time of enrollment for the OptiGene Direct Plus RT-LAMP test.

The study protocol at all the sites was approved by the respective local institutional ethical review boards.

### Test and RT-PCR assays

All the assays were performed as per the manufacturer’s instruction for use (IFU).

The Atila iAMP test was performed on the same day of test sample collection for all samples in El Salvador, and 81 of 85 (95.3%) samples in New Jersey (NJMS). The samples not tested on the same day were frozen at -20^0^C in Paraguay and -80^0^C in Wisconsin (MCW) and tested in batches. A validated RT-PCR system (i.e., Biorad CFX96 RT system or Atila PowerGene 9600 Plus RT-PCR system) with FAM/HEX fluorescence detection was used for reaction run and detection. Positive and negative controls were run for each batch, and the batch was considered valid only if both controls were valid. The individual sample test result was determined to be positive if an exponential amplification curve with cycle threshold (ct)<50 was present in the FAM (ORF-1a/b or N-genes) channel. The test result was determined to be negative if the FAM channel did not have an amplication curve, and the HEX (internal control) channel had an exponential amplification curve with ct<50. The test was determined invalid if no amplification was detected in both FAM and HEX channels, in which case the test was repeated. If the repeat run was also invalid, then the that sample was considered invalid. In total, 1.0% of NP, 1.4% of anterior nares, 3.9% of mid-turbinate, and 0% of saliva samples had invalid results. Less than 1% (0.6%) of saliva samples could not be tested secondary to the samples being predominantly phlegm.

With few exceptions [5 of 85 (6.0%)], the OptiGene Direct Plus RT-LAMP test was performed on the same day of the test sample collection. A Genie^®^ III or II platform (OptiGene, UK) was used for the reaction run and detection. Positive and negative controls were run for each batch of samples, and the batch was considered valid only if both controls were valid. The Genie® software automatically analyzed the individual sample test results as positive or negative based on the amplification plot and annealing temperature. The test result is reported positive if the fluorescence level of the amplification curve rises above a defined threshold and the peak of the annealing curve is above a defined threshold and lies within a specified temperature range. All of the OP and anterior nares samples were tested. 19% of the saliva samples were not tested because those samples were predominantly phlegm without saliva.

A single run was performed for each sample at all study sites except at NJMS which performed duplicate runs for each sample. To ensure comparability across the sites, for the pooled analysis, the first of the duplicate run at NJMS was used.

### Statistical analysis

Pooled and study site-specific analyses were performed overall and stratified by different sampling strategies. For the overall analysis, if any sample anatomic collection site tested positive, that subject was identified as positive for that test assay. If all collection site samples were negative for the subject, the subject was considered negative for that test assay.

The NP sample for the RT-PCR test used for clinical diagnosis was considered the reference method. The sensitivity was defined as the proportion of RT-PCR positive samples which tested positive by the test assay, and specificity was defined as the proportion of the RT-PCR negative samples which tested negative by the test assay. Additional stratified analyses by the ct-value for the RT-PCR, as a surrogate marker for the viral load, and history of symptoms were also performed wherever the data was available. History of symptoms was collected from the subjects at the time of sample collection. 95% confidence intervals (CI) were calculated for the sensitivity and specificity measures. Imbalances in paired sample results were evaluated using Mc-Nemar’s test, with a p-value <0.05 considered statistically significant. Data analysis was performed using IBM^®^ SPSS software.

## Results

### Atila iAMP test

In the overall analysis (Figure 1), the sensitivity of the Atila iAMP test was 76.2% (95% CI: 71.1-80.7) and the specificity was 94.9% (95% CI: 93.3-96.1) for detection of SARS-CoV-2. Stratified by study site, the sensitivity was 63.8% (95% CI: 55.9-71.2) in El Salvador, 88.5% (95% CI: 79.9-94.3) in Paraguay, 88.9% (95% CI: 65.3-98.6) in Wisconsin, and 89.1% (95% CI:77.8-95.9) in New Jersey. The specificity was 97.2% (95% CI: 95.7-98.2), 81.3% (95% CI: 74.7-86.7), 100% (95% CI: 96.6-100) and 100% (95% CI: 88.4-100), respectively. Since the El Salvador site’s sensitivity was significantly lower than all the other sites, and considered an outlier, we conducted an overall pooled analysis excluding El Salvador, which demonstrated an overall sensitivity of 88.8% (95% CI: 82.8-93.2) and an overall specificity of 89.5% (95% CI: 85.6-92.7).

**Figure 1:**
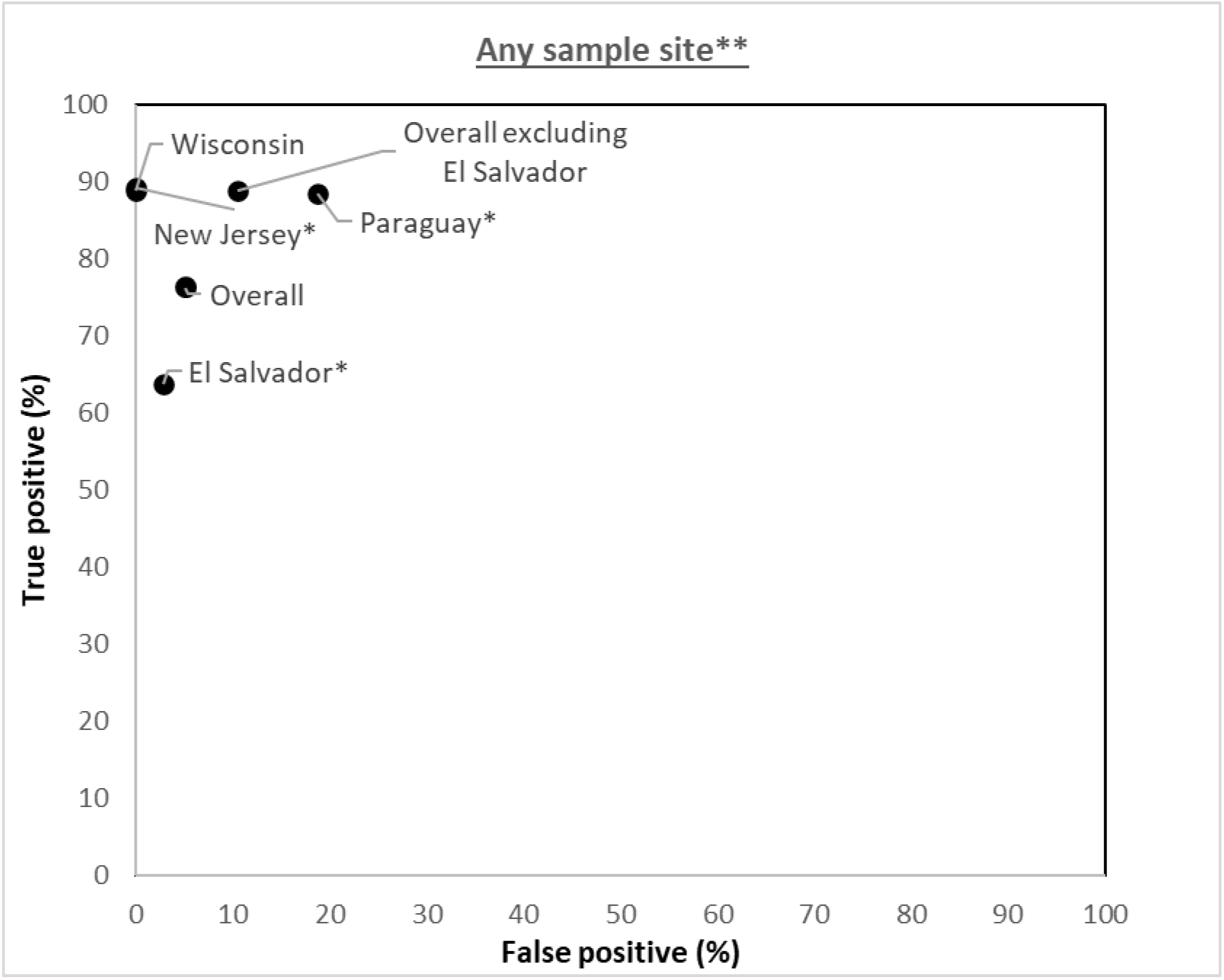
Study site specific analysis of validity of Atila iAMP assay against PCR (Reference) test (not stratified by sample collection site) ^*^P-value < 0.05 for McNemar’s test (continuity corrected); **Any sample collection site positive out of the total samples collected is considered positive

We evaluated the clinical performance of individual sampling strategies (Figure 2). The sensitivity and specificity of the provider-collected NP sample was 65.4% (95% CI: 59.9-70.6) and 97.6% (95% CI: 96.5-98.4). Since sensitivity at the El Salvador site was significantly different than all the other sites, we recalculated the overall sensitivity excluding El Salvador, which led to the sensitivity of 78.9% (95% CI: 71.6-85.1) and specificity of 95.4% (95% CI: 92.4-97.5).

**Figure 2:**
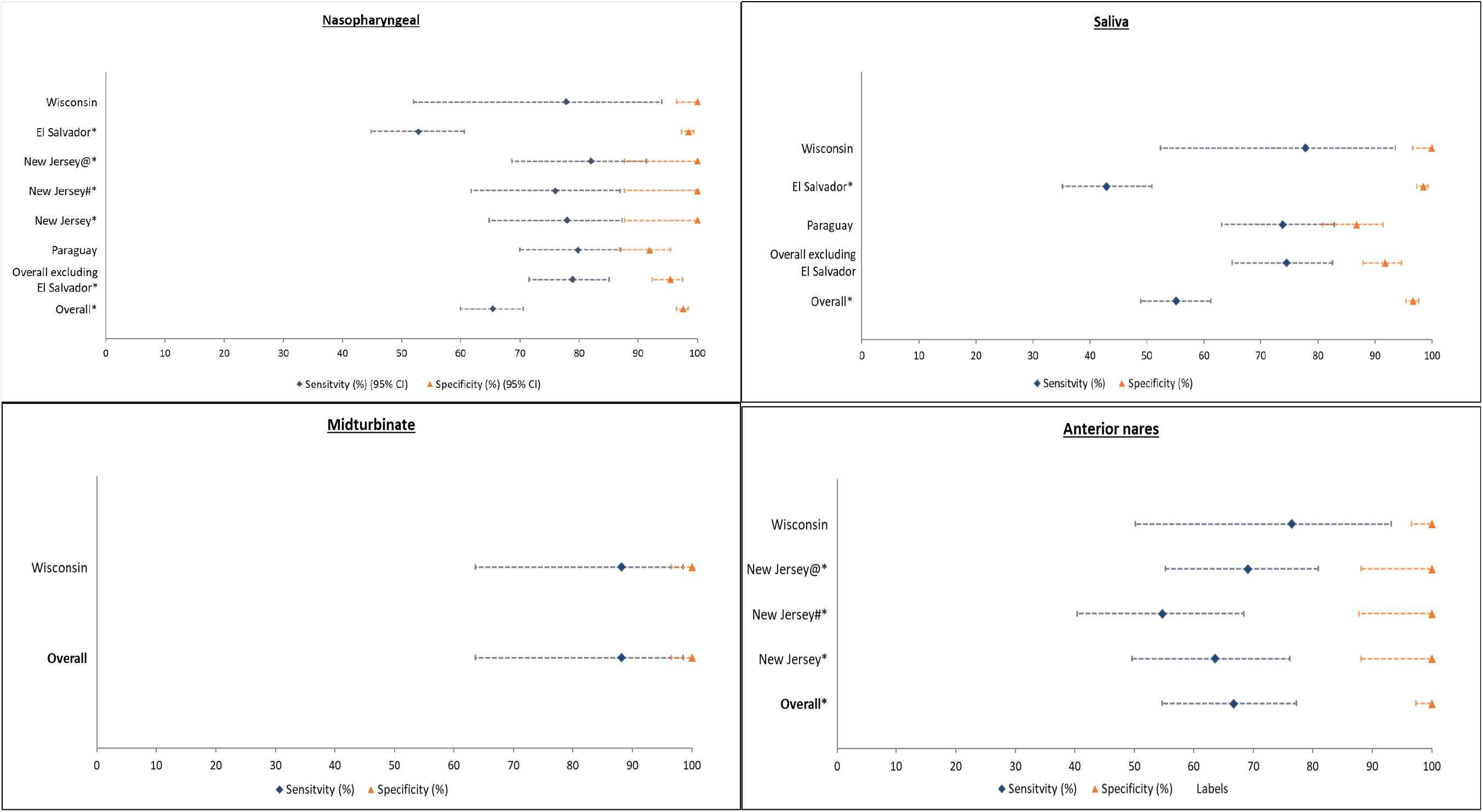
Study site specific analysis of validity of Atila iAMP assay against PCR (Reference) test (stratified by sample collection site) ^*^P-value < 0.05 for McNemar’s test (continuity corrected); #Samples were tested in duplicates and the test was considered positive only if both were positive; @Samples were tested in duplicates and the test was considered positive if either was positive

Comparing the other sampling strategies to the reference standard NP sample, self-collected dry mid-turbinate sample (only collected at MCW) was found to be most sensitive [88.2% (95% CI: 63.6-98.5)] and specific [100% (95% CI: 96.5-100)]. Self-collected saliva samples, excluding El Salvador (due to significantly different estimate than other sites), had an overall sensitivity of 74.5% (95% CI: 64.9-82.6) and overall specificity of 91.8% (95% CI: 87.9-94.7). The self-collected dry anterior nares sample was the least sensitive strategy with the overall sensitivity of 66.7% (95% CI: 54.6-77.3) and overall specificity of 100% (95% CI: 97.3-100). Since anterior nares sample was not collected at the El Salvador study site and none of the study sites had significantly different estimate than other sites for anterior nares sample, no exclusion was made.

Assuming that viral load would influence accuracy, we analyzed the sensitivity at different ct-values on RT-PCR among the positive subjects (Figure 3). Restricting the analysis to samples with ct≤35, ct≤30, ct≤25, and ct≤20 increased the sensitivity to 82.6%, 97%, 100% and 100% for NP samples and 68%, 86.1%, 88.9%, and 100% for anterior nares samples in New Jersey (NJMS) and 79.8%, 81.3%, 88.4%, and 100% for NP samples and 73.8%, 73.8%, 79.7%, and 84.3% for saliva samples in Paraguay. The respective corresponding percentages for El Salvador were 65.9%, 78.1%, 83.6%, and 89.1% for NP samples and 52.8%, 58.3%, 64.4% and 65.5% for saliva samples. Although the sensitivity increased in El Salvador with an increase in viral load (i.e, at lower ct-values), within each ct-value strata, the sensitivity in El Salvador was still lower than in Paraguay and New Jersey for each anatomic collection sites.

**Figure 3:**
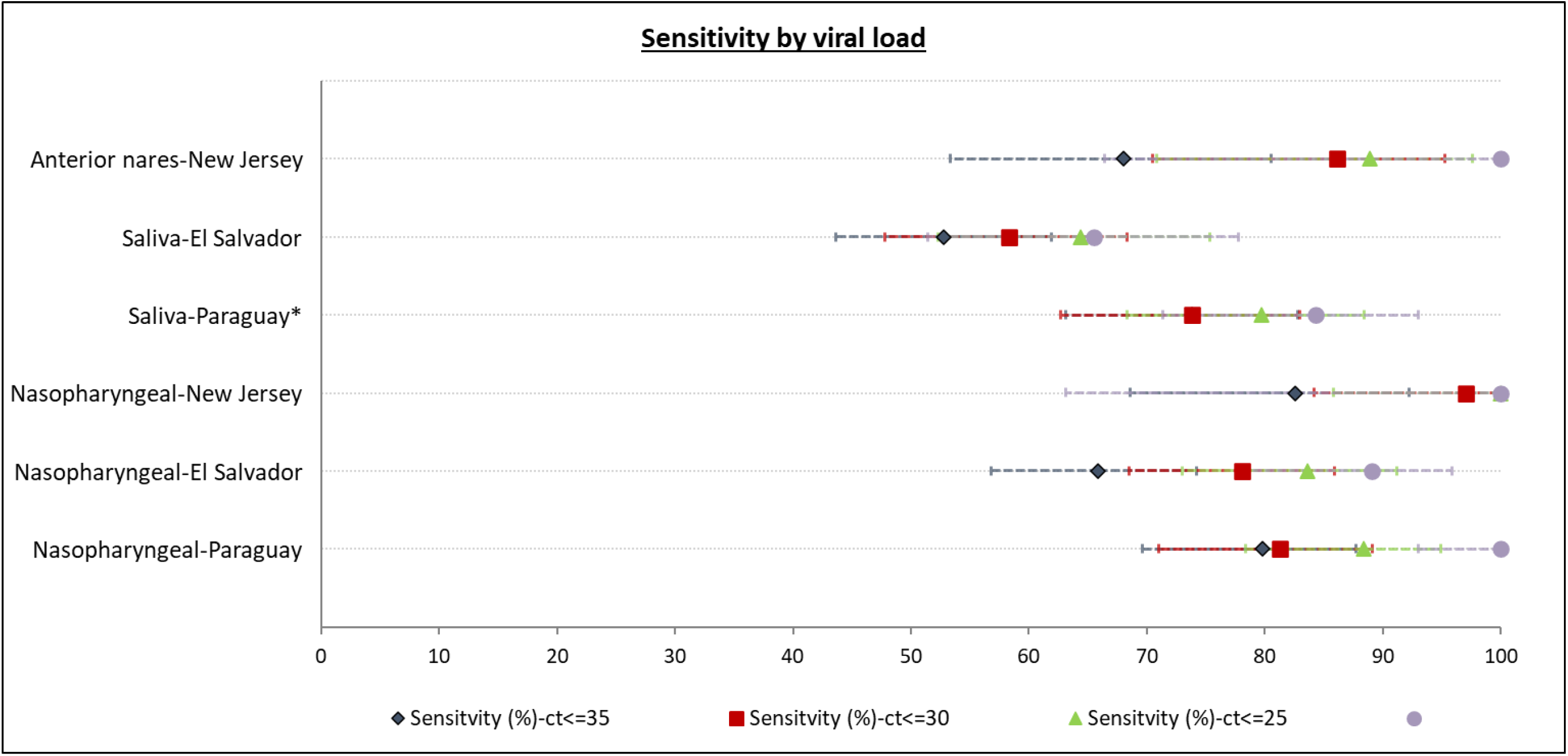
Study site and sample site specific analysis of the sensitivity of Atila iAMP assay against PCR (Reference) test stratified by the ct-values. ^*^Sensitivity for ct<35 and ct<30 was equal

The mean ct-value on RT-PCR in El Salvador among the RT-PCR positive asymptomatic subjects was 30.0 (95% CI: 25.6-30.4), and among symptomatic subjects was 22.5 (95% CI: 20.2-24.8). The respective corresponding values in Paraguay were 18 (95% CI: 13.7-22.4) and 17.5 (95% CI: 15.8-19.2). The mean ct-value on RT-PCR in New Jersey among the RT-PCR positive subjects was 25.7 (95% CI: 23.9-27.5); all RT-PCR positive subjects were hospitalized for observation and management of COVID-19 and so likely symptomatic. The sensitivity of the NP sample in El Salvador among symptomatic subjects was significantly higher [65.1% (95% CI: 54.1-75.1)] than among asymptomatic subjects [39.0% (95% CI: 28.0-50.8)] (Figure 4). The difference was not significant between symptomatic and asymptomatic subjects for NP samples in Paraguay, saliva in El Salvador, and saliva in Paraguay. Among the symptomatics subjects, the sensitivity was significantly higher for saliva samples in Parguay [74.3% (95% CI: 62.4-84.0)] than in El Salvador [51.2% (95% CI: 40.1-62.1)]; the difference was not significant for the NP samples in Paraguay [81.7% (95% CI: 70.7-89.9)] and New Jersey [78% (95% CI: 64-88.5)] and in El Salvador [65.1% (95% CI: 54.1-75.1)]. Among the asymptomatic subjects, the difference for either NP [69.2% (95% CI: 38.6-90.9) in Paraguay and 39% (95% CI: 28.0-50.8) in El Salvador] or saliva samples [71.4% (95% CI: 41.9-91.6) in Paraguay and 33.8% (95% CI: 23.4-45.4) in El Salvador] was not significantly different between Parguay and El Salvador.

**Figure 4:**
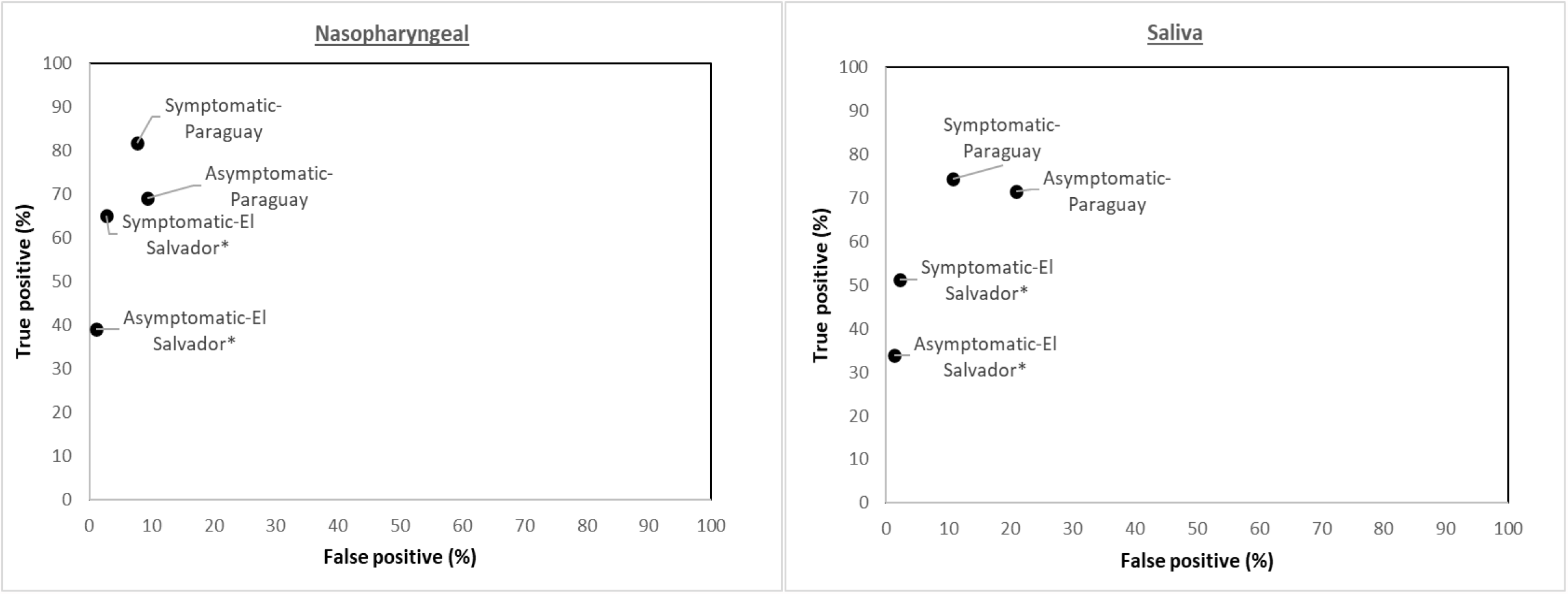
Study site and sample site specific analysis of validity of Atila iAMP assay against PCR (Reference) test stratified by the Symptoms. ^*^P-value < 0.05 for McNemar’s test (continuity corrected)

### OptiGene Direct Plus RT-LAMP test

The overall sensitivity and specificity of the OptiGene Direct Plus RT-LAMP test were 25.5% (95% CI: 14.7-39) and 100% (95% CI: 88.4-100), respectively (Figure 5). The estimates did not differ significantly by different sampling strategies or duplicate testing. Furthermore, when limiting the analysis to test samples collected within 24 hrs of RT-PCR sample collection, the overall sensitivity was still only 33.3%.

**Figure 5:**
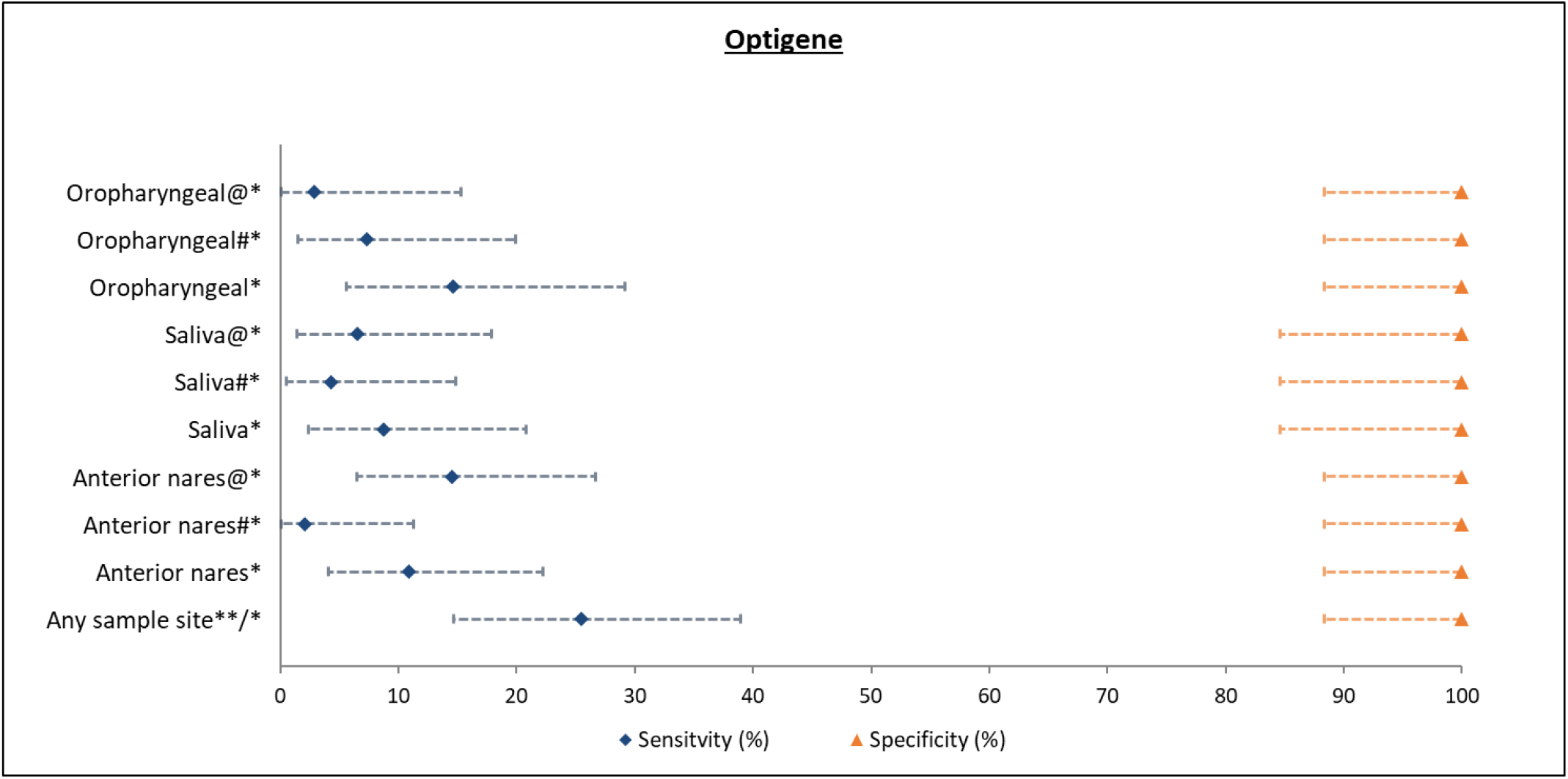
Study site specific analysis of validity of OptiGene Direct Plus RT-LAMP assay against PCR (Reference) test (overall and stratified by sample collection site) ^*^P-value < 0.05 for McNemar’s test (continuity corrected); **Any sample collection site positive out of the total samples collected is considered positive; #Samples were tested in duplicates and the test was considered positive only if both were positive; @Samples were tested in duplicates and the test was considered positive if either was positive

## Discussion

The current study evaluated the clinical performance of two isothermal amplification tests for detection of SARS-CoV-2. The overall sensitivity and specificity of the Atila iAMP test for detection of SARS-CoV-2, excluding the outlier study site, were 88.8% and 89.5%, respectively. The sensitivity, excluding the outlier study site, was 78.9% for nasopharyngeal, 88.2% for self-sampled mid-turbinate, 74.5% for direct saliva and 66.7% for anterior nares samples. The specificity for these sites ranged from 91.8% to 100%. The sensitivity increased with higher viral load (i.e., at lower ct-values) and among symptomatic as compared to asymptomatic participants. The sensitivity and specificity of the OptiGene Direct Plus RT-LAMP test, conducted at a single site, were 25.5% and 100%, respectively.

There is scant literature on the performance of the Atila iAMP COVID test. We identified only one clinical performance evaluation of the Atila iAMP COVID test on the direct, non-extracted samples, which is the recommended application as per EUA by the manufacturer. This small-scale evaluation (n=197) showed a sensitivity of 44.1% and specificity of 96.6% for the Atila iAMP test on NP swabs with a large number (35.5% or 70/197) of invalid results (12). A small (n=50) analytic and clinical validation study on the Atila iAMP assay showed the analytic LOD for the assay to be 50-100 copies/reaction for ORF1-a/b gene and 1-10 for the N gene, which is higher than that of RT-PCR (average range of 1-10) (13). This may explain our finding of lower clinical sensitivity of the assay at higher ct-values, considering ct-values as a surrogate marker for the viral load, which may not always be precise (14). In the clinical validation by the same group, the assay was found to have 100% agreement with the RT-PCR. However, this validation was on extracted RNA and was based on 46/50 samples that have ct≤30. Another small (n=50) clinical validation (15), again on extracted RNA, showed the sensitivity and specificity of the assay to be 82.8% and 100%, respectively, with all five false-negative samples to have ct≥35.

The OptiGene Direct Plus RT-LAMP COVID assay has been previously clinically validated by the NHS trust to have a sensitivity of 70% for swabs and 79% for saliva, with an increase in sensitivity to 100% for swabs at ct≤25 (16). However, similar to our validation, such high sensitivity was not confirmed by other groups, which showed the sensitivity in the range of 46.7% (17)and 34%, including false-negative results on symptomatic high viral load subjects (18). Our validation study was based on kits purchased from the manufacturer, using fresh samples (not freeze-thawed samples) collected and placed in the VTM recommended by the manufacturer, and run as per the instructions provided by the manufacturer. Furthermore, even though the reference RT-PCR used in our assay targeted E or N2 and S gene in addition to the ORF1-a/b gene, given that a NP swab based RT-PCR is the accepted reference standard for the SARS-COV-2 diagnosis (7,8), we believe that clinical sensitivity of the assay should not be affected by the differences in gene targets between the assays. While it has been suggested that assays targeting the N gene are not a valid reference standard to evaluate the OptiGene Direct Plus RT-LAMP assay (19), this is not supported by clinical studies.

It is important to note that the IFU’s for both assays state the need to confirm the negative test result with a more sensitive RT-PCR test, and do not claim to be the final screening answer (20). However, as compared to the RT-PCR assays, which may sometimes take >24 hr of turnaround time (TOT) with considerable cost, the isothermal amplification-based assay’s advantage is its rapid TOT (∼1 hour), lower cost, and ease of performance (no nuleic acid extraction needed).

Thus, it can cheaply and rapidly identify high viral load subjects who are likely to be most infectious (21)(22). Moreover, there is at least some evidence to suggest that RT-PCR positivity does not necessarily translate into infectivity because it can detect the shedding of post-infectious viral RNA particles shedding, particularly among post-symptomatic patients (23,24). The Atila iAMP has similar advantages as the rapid antigen tests with regard to ease of operability and quick TOT, but provides higher sensitivity (reported to be 67-73% for rapid antigen test (25,26)) resulting in more reassurance of a negative test result.

Variation in the performance of both the assays across various study sites in our evaluation and notable differences to other studies cannot be ignored. It demonstrates the limitations of EUAs which may not necessarily translate to acceptable clinical performance for all tests in all settings. A thorough clinical validation of diagnostic assays on a standardized panel of samples in clinical settings is advisable before its widespread adoption for clinical use.

We do not fully understand the reason for the variation in test performance across study-sites. Importantly, the populations at each site was different with respect to SARS-CoV-2 prevalence, clinical symptoms, and other factors, but stratified analyses showed similar performance at all sites except for El Salvador. Given that invalid results were rare and did not differ across the study sites and there was no consistent pattern observed in ct-values for the internal control [mean ct-values for the internal control for NP: 28.4 (95% CI: 28.1-28.7) (El Salvador), 34.1 (95% CI: 32.8-35.1) (MCW), 22.7 (21.9-23.4) (Paraguay), 25.4 (23.4-27.3) (NJMS); for saliva: 19.4 (95% CI: 19.1-19.7), 23.6 (95% CI: 23.1-24.2) (MCW), 22.1 (21.3-22.8) (Paraguay)], we do not attribute the lower sensitivity in our validation in El Salvador to sampling variation. Rather we hypothesize the lower sensitivity of the Atila iAMP test in El Salvador to be related to multiple factors: relatively higher proportion of asymptomatic subjects as compared to Paraguay (69.8% versus 21.9%) and operator-dependent nature of the assay due to the hands-on nature of the test to set up the reaction (27). However, given that on stratified analysis by symptoms and ct-values, the sensitivity was still lower in El Salvador than other sites within the strata, makes the second explanation more likely. Variation in the reference standard RT-PCR method and RNA extraction kits used across the study sites as well as variation in duration of performing the test assay after collection is a limitation of our study and may also have influence on the study site-wide variations.

## Conclusions

In this first large-scale multi-site clinical evaluation of the Atila BioSystems iAMP COVID-19 detection test, the assay showed good sensitivity with high specificity for detection of SARS-CoV-2, particularly on high viral load (i.e., ct≤25) NP samples. In addition, it also showed moderate sensitivity for ct≤35 NP samples and ct≤25 saliva samples. Overall, the sensitivity was superior for NP and mid-turbinate samples compared to saliva and anterior nares samples. The rapid TOT, low cost, and lack of need for nucleic acid extraction make Atila iAMP test a reasonable alternative screening test for SARS-COV-2 for patients and providers in outpatient clinics to identify likely infectious subjects. When implemented with other COVID safety measures, such low cost testing can provide an approach for the safe reopening and daily clinical activities of essential medical services for the highest risk population in immediate need of care. However, inconsistency observed in assay performance across the study sites highlights the need for a rigorous site-specific clinical performance evaluation of the isothermal-amplification-based assays before their clinical adoption.

## Data Availability

The datasets used in the current study are available from the corresponding author on reasonable request.

## Acknowledgements

The field effort was a collaboration of the US National Cancer Institute (NCI) with Basic Health International (BHI), USA and Rutgers New Jersey Medical School (NJMS), USA. The BHI had collaboration with Hospital Nacional de Santa Ana, El Salvador, Hospital Materno Infantil de San Lorenzo, Ministerio de Salud Pública y Binestar Social (MSP-BS), Paraguay, Centro de Especialidades Dermatológicas, MSP-BS, Paraguay, Instituto de Investigaciones en Ciencias de la Salud, Universidad Nacional de Asunción, Paraguay,, and Medical College of Wisconsin, USA.

## Author’s contributions

NW, APN, NCD, JF, MM, RM, MC, KTD contributed substantially to the conception and design of the study. KA, LM, MM, RD, AV, CDA, MM, JF, MP, APN, MHE contributed to acquisition of data. MF, SG, NCD, BM contributed to running the test assays. KTD, LM, NW contributed to the analysis and interpretation. KTD, NW drafted the manuscript. All authors provided critical revision of the article and provided final approval of the version to publish.

## Ethical approval and informed consent

The study was approved by the ethical review board of Comite Nacional de Etica de Investigacion en Salud (IRB no.FWA00010986) in El Salvador, Comité de Ética, Instituto de Investigaciones en Ciencias de la Salud, Universida Nacional de Asución (IRB no. P37/2020) in Paraguay, MCW IRB (IRB no. FWA00000820) in Wisconsin, and the Newark Health Sciences IRB for Rutgers Biomedical Health Sciences (IRB no. Pro2020001801) in New Jersey. Written informed consent was obtained from all study participants.

## Abbreviations

CI: Confidence Interval
COVID-19: Coronavirus Disease of 2019
CT: Cycle Threshold
EUA: Emergency Use Authorization
iAMP: Isothermal Amplification
IFU: Instruction for Use
LAMP: Loop-mediated Isothermal Amplification
LMIC: Low- and Middle-Income Countries
LOD: Limit of Detection
MCW: Medical College of Wisconsin
NAAT: Nucleic Acid Amplification Test
NP: Nasopharyngeal
OP: Oropharyngeal
NJMS: Rutgers New Jersey Medical School
RT-PCR: Reverse Transcription-Polymerase Chain Reaction
SARS-COV-2: Severe Acute Respiratory Syndrome Coronavirus 2
SPSS: Statistical Package of Social Studies
VTM: Viral Transport Medium

